# Automated detection of large vessel occlusion: a multicenter study validating efficacy and proving clinical implications

**DOI:** 10.1101/2024.05.08.24307038

**Authors:** Kyu Sun Yum, Jong-Won Chung, Sueyoung Ha, Kwang-Yeol Park, Dong-Ick Shin, Hong-Kyun Park, Yong-Jin Cho, Keun-Sik Hong, Jae Guk Kim, Soo Joo Lee, Joon-Tae Kim, Oh Young Bang, Myungjae Lee, Dong-Min Kim, Leonard Sunwoo, Hee-Joon Bae, Wi-Sun Ryu, Beom Joon Kim

## Abstract

**Objective:** We aimed to validate a software, JLK-LVO, that automatically detects large vessel occlusion (LVO) on computed tomography angiography (CTA) using deep learning, within a prospective multicenter dataset. In addition, we calibrated the predicted probability of LVO against observed frequency and assessed the clinical implications of LVO probability in terms of follow-up infarct volume and functional outcome.

**Method:** From 2021 to 2023, we prospectively collected data from patients who underwent CTA within 24 hours of symptom onset at six university hospitals in Korea. The diagnostic performance of the software was evaluated using the area under the curve (AUC), sensitivity, and specificity across the entire study population and specifically in patients with isolated middle cerebral artery (MCA)-M2 occlusion. In addition, we compared LVO probabilities after stratifying patient into acute LVO, chronic LVO, isolated MCA-M2 occlusion, relevant MCA stenosis, and without steno-occlusion of MCA groups. We calibrated LVO probabilities in two ways: through mathematical calibration using logistic regression, and by refining LVO probabilities based on the observed frequency of LVO. We then assessed the association of LVO probability categories with infarct volume on follow-up diffusion-weighted imaging (DWI) and modified Rankin Scale (mRS) scores three months post-stroke, using ANOVA and the Cochran–Armitage test.

**Results:** After excluding 168 patients, 796 remained; the mean (SD) age was 68.9 (13.7) years, and 57.7% were men. LVO was present in 193 (24.3%) of these patients, and the median interval from last known well to CTA was 5.7 hours (IQR 2.5 to 12.1 hours). At default threshold of 0.5, the software achieved an AUC of 0.944 (95% CI 0.926–0.960), with a sensitivity of 0.896 (0.845–0.936) and a specificity of 0.904 (0.877–0.926). In isolated MCA-M2 occlusion, the AUROC was 0.880 (95% CI 0.824–0.921). Compared to the without steno-occlusion of MCA groups (median LVO probability 0.5, interquartile range 0.1 – 6.5), relevant stenosis (median 15.3, 2.4 –77.4) and isolated MCA-M2 occlusion (82.1, 40.9 – 98.2) groups had significantly higher LVO probability. Due to sparse data between 20-60% of LVO probabilities, recategorization into unlikely (0-20% LVO scores), less likely (20-60%), possible (60-90%), and suggestive (90-100%) provided a reliable estimation of LVO compared with mathematical calibration. The category of LVO probabilities was significantly associated with follow-up infarct volumes on DWI and 3-month mRS scores.

**Conclusion:** In this multicenter validation study, we proved the clinical efficacy of the software in detecting LVO on CTA. Additionally, using large-scale real-world data, we calibrated the LVO probabilities, which may provide a more confident estimation of LVO for practicing physicians.

## Introduction

Recent advancements in stroke imaging and the development of procedural devices have extended the therapeutic window for endovascular therapy (EVT) in patients with large vessel occlusion (LVO).^1^ Accrued evidence has redefined the standard of care for LVO patients who present within 6 to 24 hours from their last known well time.^2,3^ These trials predominantly depend on magnetic resonance (MR) perfusion or computed tomography (CT) perfusion imaging to determine clinical or tissue mismatches.^2,3^ Despite the critical role of these advanced imaging modalities, their availability remains limited at most primary stroke centers globally.^4^ Recent research has advocated for the adoption of more readily available imaging techniques, such as CT angiography (CTA). Notably, the CT for Late Endovascular Reperfusion (CLEAR) trial demonstrated that clinical outcomes for patients assessed using non-contrast CT alongside CTA were on par with those evaluated using CT perfusion or MR perfusion.^5^ Additionally, a sub-study by the HERMES collaboration (Highly Effective Reperfusion Evaluated in Multiple Endovascular Stroke Trials) within the early time window (0-6 hours) indicated comparable rates of favorable functional outcomes between patients undergoing CT perfusion and those who did not.^6^

Initially, 66% of EVT candidates route to centers lacking the capability to perform EVT,^7^ despite better prognoses at facilities equipped for such treatments. Thus, it is crucial for centers not capable of EVT to accurately and promptly identify LVO at any time—around the clock—to ensure rapid patient transfer to EVT-capable centers. However, the lack of vascular specialists is a significant barrier for many smaller hospitals. Even within EVT-capable centers, enhancing the ability to screen CTA for LVO could improve procedural efficiency, staffing, and reduce the time from patient arrival to treatment initiation.

Automated LVO detection software (JLK-LVO, JLK Inc., Korea), which uses a deep learning algorithm, presumably utilizes the symmetry of vessel density between hemispheres. Since vessel density may reflect perfusion status,^8^ the deep learning-derived LVO probability score—primarily based on comparisons of vessel density between hemispheres—could be associated with clinical outcomes following ischemic stroke, particularly in terms of follow-up infarct volumes and functional outcomes. Furthermore, due to the nature of deep learning, more accurate models produce poorly calibrated predictions.^9^ Hence, the probability derived from the algorithm needs to be calibrated using real-world observations, which has been less highlighted in the application of deep learning algorithms in the medical field.^10^

In this prospective multicenter study, we aimed to clinically validate the automated LVO detection software and to calibrate the probability of the deep learning algorithm using real-world data. In addition, we sought to investigate the clinical implications of calibrated LVO probability scores in relation to infarct volumes on follow-up diffusion-weighted imaging and functional outcomes 3 months post-ischemic stroke.

## Methods

### Study populations

We consecutively enrolled patients with acute ischemic stroke or transient ischemic attack who were admitted to participating stroke centers. Enrollment occurred from April 2022 to April 2023 at five centers and from January 2021 to March 2022 at one center (Supplementary Figure 1). Exclusion criteria were 1) CTA performed after 24 hours of symptom onset, 2) poor image quality of insufficient contrast to analyze, and 3) hemorrhagic transformation or brain tumor. All patients or their legal representatives gave a written informed consent. The study protocol was approved by institutional review board of Seoul National University Bundang Hospital [B-2307-841-303].

### Clinical data collection

We retrieved baseline demographic and clinical information for all study participants from a web-based prospective stroke cohort (strokedb.or.kr), including age, sex, history of previous stroke, functional status before stroke, and cardiovascular risk factors such as hypertension, diabetes mellitus, habitual smoking, and atrial fibrillation. The stroke characteristics included the time interval between the onset of symptoms and time of CTA, the National Institutes of Health Stroke Scale (NIHSS) score at admission, and treatment information. The functional status at 3 months after stroke was measured using the mRS score, which was determined through a structured telephone interview by an experienced physician assistant at each hospital.

### Brain imaging

CT angiography images were acquired according to standard departmental protocols in each hospital. The scanning parameters were 90∼120 kVp, 60∼376 mAs, 38.4 or 40-mm beam collimation, 0.33∼0.6-second rotation time, and 0.625 ∼ 3-mm section thickness (Supplementary Table 1). Diffusion-weighted images were acquired using 1.5 or 3.0 T MRI systems (majority [>95%] of systems are Phillips or Siemens). Slice thickness was 3–5 mm, spacing between slices 3.3–6.5 mm, pixel spacing 0.469 – 1.375 mm, repetition time 2426 – 8800 ms, echo time 64 – 108 ms.

### Definition of large vessel occlusion

In the present study, large vessel occlusion (LVO) was operationally defined as an arterial occlusion encompassing the intracranial segment of the internal carotid artery (ICA), as well as the M1 and M2 segments of the middle cerebral artery (MCA-M1 and MCA-M2, respectively). The term “intracranial ICA” specifically denotes the segment extending from the petrous part to the bifurcation with the MCA and the anterior cerebral artery (ACA).^11^ The MCA-M1 segment encompasses the stretch from the MCA-ACA bifurcation to the initial branching of the MCA, while the MCA-M2 segment includes the part ascending vertically along the Sylvian fissure from the MCA branching point.^11^ In cases where the MCA divided early, a functional classification was utilized whereby the segment closest to the origin was labeled as M1, with subsequent downstream branches classified as M2.^12^ To confirm the presence of LVO, CTA source images, maximum intensity projection (MIP) images, and three-dimensional rendering images were thoroughly examined by two experienced vascular neurologists (W-S. R and S.H), alongside an evaluation of patients’ magnetic resonance imaging (MRI) scans and symptomatic data. In cases of diagnostic discrepancy, a final determination was made by an experienced neuroradiologist (L. S). Along with the presence of LVO, location (ICA, MCA-M1, and MCA-M2), and the side of LVO were recorded. We define acute LVO as LVO relevant to the index stroke, whereas chronic LVO is defined as LVO that is not relevant to the index stroke. Relevant MCA stenosis is defined as moderate to severe stenosis on CTA that corresponds to infarcts observed on DWIs.

### Deep learning-based software

Source images of CTA with slice thickness between 0.5 – 2 mm were fed into the commercially available deep learning-based software (JLK-LVO, JLK Inc., Seoul, Korea).^13^ In brief, an automated algorithm selects slices from source images to construct MIP images. The vessel segmentation involves a 2D U-Net based on the Inception Module,^14^ trained to segment vessels in axial MIP images. A vessel occlusion detection algorithm follows, involving the combination of vessel masks into a compressed image for training an EfficientNetV2 model.^15^ Finally, the model produced LVO score, probability of LVO by the algorithm, and the side of LVO based on comparison of heatmap size between hemispheres.

### Image analysis

Infarct location was categorized as anterior circulation, posterior circulation, and multiple based on the review of follow-up DWI by an experienced vascular neurologist (J-W. Chung). Follow-up diffusion-weighted images within 7 days after CTA were included to analyze the association between LVO score and follow-up infarct volumes. Infarct volumes on DWI were calculated using a validated software package (JLK-DWI, JLK Inc., Seoul, Korea).^16,17^ Segmented infarct area was meticulously supervised by an experienced vascular neurologist (W-S. Ryu).

### Statistical analysis

Baseline characteristics among participating centers were compared using the ANOVA or Kruskal-Wallis test for continuous variables, and the chi-square test for categorical variables, as appropriate. To validate the accuracy of the software in diagnosing LVO, we computed the AUROC, as well as sensitivity, specificity, PPV, and NPV. A 1000-repeat bootstrap analysis was employed to calculate the 95% confidence intervals (CIs) for all parameters. The AUROC was used in combination with the DeLong method^18^ to compute the standard error (SE) of the AUROC. The cutoff for the LVO score used in the analysis was set at 0.5. A true positive was defined when both the presence and side of LVO were concordant between JLK-LVO and the experts’ consensus. If the presence of LVO was correctly identified but the side was incorrect, the case was classified as a false negative. We conducted additional analyses to determine the optimal threshold that would yield the maximum Youden index (sensitivity + specificity −1). Given that the software is primarily intended for screening LVO, we also computed specificity, PPV, and NPV at a sensitivity level of 0.90. Furthermore, we performed the AUROC analysis at each participating center. To test the deep learning algorithm’s ability to detect isolated MCA-M2 occlusion, we reran the analysis for patients with isolated MCA-M2 occlusion, including those without LVO as the control group. After stratifying patients into groups—acute LVO, chronic LVO, isolated MCA-M2 occlusion, relevant MCA stenosis, and neither stenosis nor occlusion of MCA—we compared LVO scores using ANOVA with Tukey for multiple comparison. We calibrated the LVO probability score using real-world data in two ways. First, we ran a logistic regression model where the ground truth label of LVO was entered as the dependent variable, and we then acquired adjusted probabilities. Using the ‘pmcalplot’ command in STATA,^19^ we displayed a calibration plot comparing observed to expected probabilities, using either unadjusted or adjusted probability scores. Second, we divided patients into ten groups at 10% intervals of LVO probability and calculated the observed frequency of LVO in each group. Subsequently, we arbitrarily categorized patients into four groups based on the observed frequency of LVO. The association between the calibrated LVO groups and infarct volumes on DWI was analyzed using dot plots and ANOVA with Tukey post-hoc comparison. Additionally, the relationship between the calibrated LVO groups and the 3-month mRS score was analyzed using the Cochran–Armitage test. All statistical analyses were performed using STATA software (version 16.0, TX, USA) and MedCalc (version 17.2, MedCalc Software, Ostend, Belgium, 2017). A P value < 0.05 was considered statistically significant.

## Results

### Study population

During the study period, a total of 1,391 patients with ischemic stroke or transient ischemic attack were admitted, and 964 (69.3%) underwent CTA in the emergency room. According to the exclusion criteria, 168 patients were excluded, leaving 796 for analysis. The mean age ± SD of the study population was 68.9 ± 13.7 years, and 57.7% were male. LVO was found in 193 (24.3%) patients, and the median interval from last known well to CTA was 5.7 hours (IQR 2.5 to 12.1 hours). Demographic characteristics were comparable among the participating centers, except for a history of previous stroke (Table 1). However, the intervals from last known well to CTA, the prevalence of revascularization therapy, and infarct volumes on follow-up DWI were significantly different. Additionally, CT vendors and parameters of CTA varied significantly across participating centers (Supplementary Table 1).

**Table 1.**
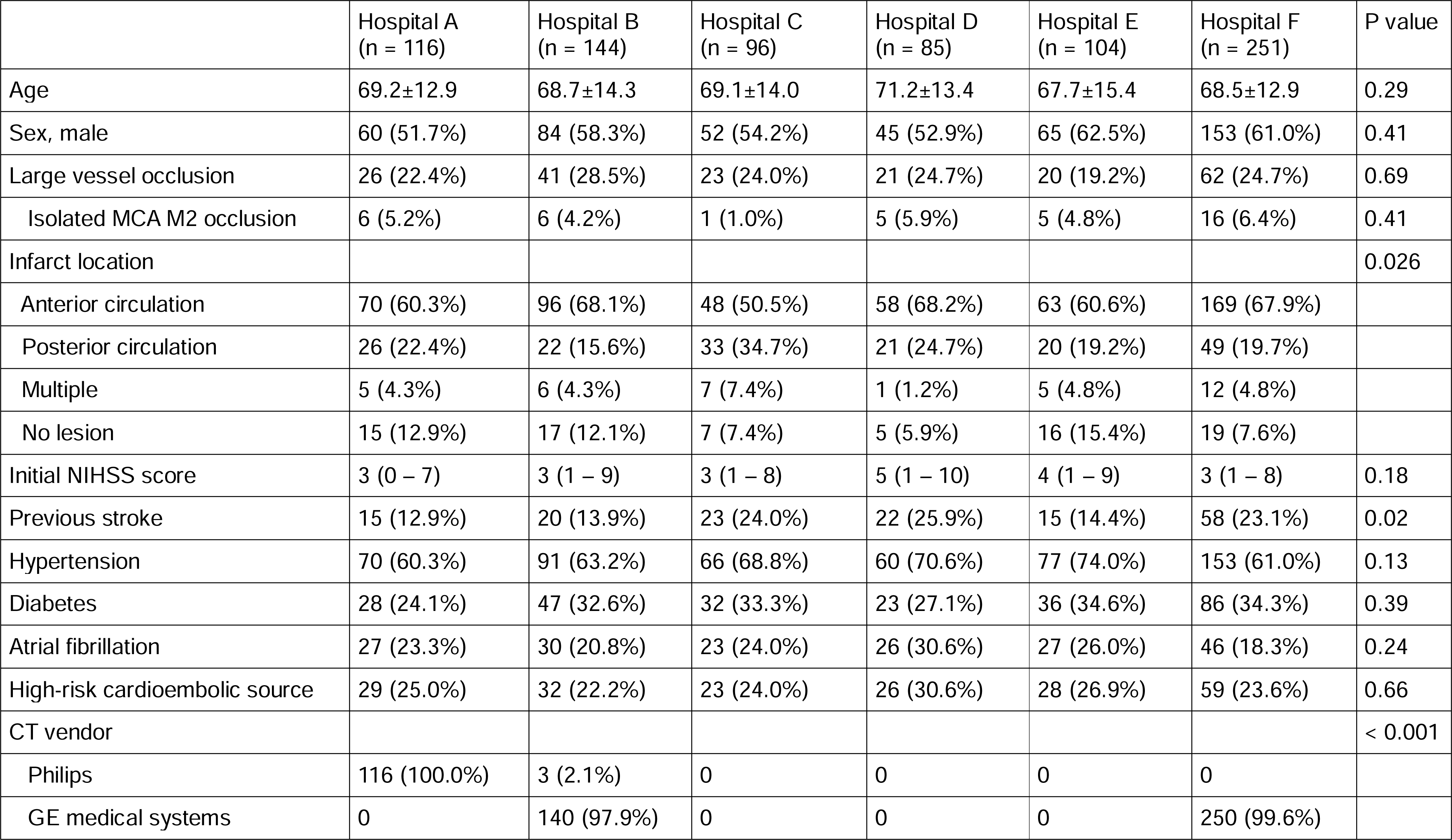
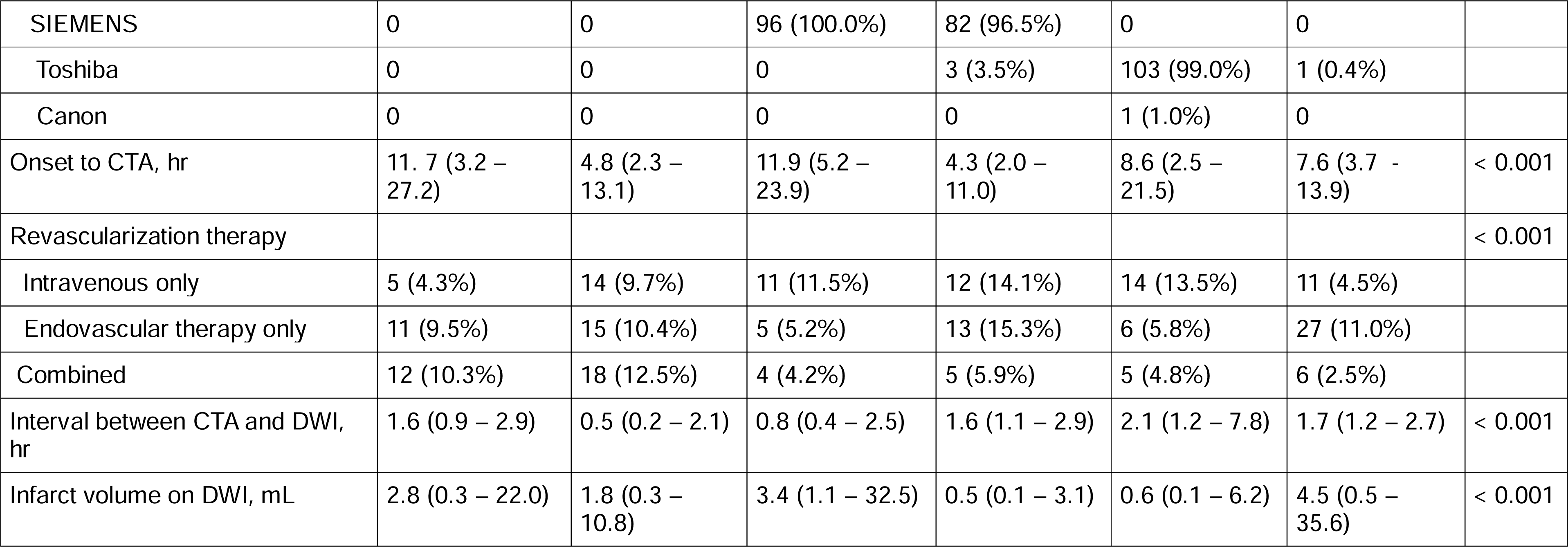
Baseline characteristics of patients in participating centers.

|  | Hospital A<br>(n = 116) | Hospital B<br>(n = 144) | Hospital C<br>(n = 96) | Hospital D<br>(n = 85) | Hospital E<br>(n = 104) | Hospital F<br>(n = 251) | P value |
| --- | --- | --- | --- | --- | --- | --- | --- |
| Age | 69.2±12.9 | 68.7±14.3 | 69.1±14.0 | 71.2±13.4 | 67.7±15.4 | 68.5±12.9 | 0.29 |
| Sex, male | 60 (51.7%) | 84 (58.3%) | 52 (54.2%) | 45 (52.9%) | 65 (62.5%) | 153 (61.0%) | 0.41 |
| Large vessel occlusion | 26 (22.4%) | 41 (28.5%) | 23 (24.0%) | 21 (24.7%) | 20 (19.2%) | 62 (24.7%) | 0.69 |
| Isolated MCA M2 occlusion | 6 (5.2%) | 6 (4.2%) | 1 (1.0%) | 5 (5.9%) | 5 (4.8%) | 16 (6.4%) | 0.41 |
| Infarct location |  |  |  |  |  |  | 0.026 |
| Anterior circulation | 70 (60.3%) | 96 (68.1%) | 48 (50.5%) | 58 (68.2%) | 63 (60.6%) | 169 (67.9%) |  |
| Posterior circulation | 26 (22.4%) | 22 (15.6%) | 33 (34.7%) | 21 (24.7%) | 20 (19.2%) | 49 (19.7%) |  |
| Multiple | 5 (4.3%) | 6 (4.3%) | 7 (7.4%) | 1 (1.2%) | 5 (4.8%) | 12 (4.8%) |  |
| No lesion | 15 (12.9%) | 17 (12.1%) | 7 (7.4%) | 5 (5.9%) | 16 (15.4%) | 19 (7.6%) |  |
| Initial NIHSS score | 3 (0 – 7) | 3 (1 – 9) | 3 (1 – 8) | 5 (1 – 10) | 4 (1 – 9) | 3 (1 – 8) | 0.18 |
| Previous stroke | 15 (12.9%) | 20 (13.9%) | 23 (24.0%) | 22 (25.9%) | 15 (14.4%) | 58 (23.1%) | 0.02 |
| Hypertension | 70 (60.3%) | 91 (63.2%) | 66 (68.8%) | 60 (70.6%) | 77 (74.0%) | 153 (61.0%) | 0.13 |
| Diabetes | 28 (24.1%) | 47 (32.6%) | 32 (33.3%) | 23 (27.1%) | 36 (34.6%) | 86 (34.3%) | 0.39 |
| Atrial fibrillation | 27 (23.3%) | 30 (20.8%) | 23 (24.0%) | 26 (30.6%) | 27 (26.0%) | 46 (18.3%) | 0.24 |
| High-risk cardioembolic source | 29 (25.0%) | 32 (22.2%) | 23 (24.0%) | 26 (30.6%) | 28 (26.9%) | 59 (23.6%) | 0.66 |
| CT vendor |  |  |  |  |  |  | < 0.001 |
| Philips | 116 (100.0%) | 3 (2.1%) | 0 | 0 | 0 | 0 |  |
| GE medical systems | 0 | 140 (97.9%) | 0 | 0 | 0 | 250 (99.6%) |  |
| SIEMENS | 0 | 0 | 96 (100.0%) | 82 (96.5%) | 0 | 0 |  |
| Toshiba | 0 | 0 | 0 | 3 (3.5%) | 103 (99.0%) | 1 (0.4%) |  |
| Canon | 0 | 0 | 0 | 0 | 1 (1.0%) | 0 |  |
| Onset to CTA, hr | 11.7 (3.2 – 27.2) | 4.8 (2.3 – 13.1) | 11.9 (5.2 – 23.9) | 4.3 (2.0 – 11.0) | 8.6 (2.5 – 21.5) | 7.6 (3.7 – 13.9) | < 0.001 |
| Revascularization therapy |  |  |  |  |  |  | < 0.001 |
| Intravenous only | 5 (4.3%) | 14 (9.7%) | 11 (11.5%) | 12 (14.1%) | 14 (13.5%) | 11 (4.5%) |  |
| Endovascular therapy only | 11 (9.5%) | 15 (10.4%) | 5 (5.2%) | 13 (15.3%) | 6 (5.8%) | 27 (11.0%) |  |
| Combined | 12 (10.3%) | 18 (12.5%) | 4 (4.2%) | 5 (5.9%) | 5 (4.8%) | 6 (2.5%) |  |
| Interval between CTA and DWI, hr | 1.6 (0.9 – 2.9) | 0.5 (0.2 – 2.1) | 0.8 (0.4 – 2.5) | 1.6 (1.1 – 2.9) | 2.1 (1.2 – 7.8) | 1.7 (1.2 – 2.7) | < 0.001 |
| Infarct volume on DWI, mL | 2.8 (0.3 – 22.0) | 1.8 (0.3 – 10.8) | 3.4 (1.1 – 32.5) | 0.5 (0.1 – 3.1) | 0.6 (0.1 – 6.2) | 4.5 (0.5 – 35.6) | < 0.001 |

### Performance of JLK-LVO

Histogram of LVO score stratified by the presence of LVO showed that the algorithm clearly differentiates LVO from non-LVO (Supplementary Figure 2). The software achieved an AUROC of 0.944 (95% CI, 0.926 – 0.960; Figure 1A) at a cutoff point of 0.50 in the entire population. The sensitivity, specificity, PPV, and NPV were 0.896, 0.904, 0.749, and 0.965, respectively (Table 2). The highest Youden index was observed at the optimal cutoff point of 0.405, with corresponding values of 0.912 for sensitivity, 0.894 for specificity, respectively. At a given sensitivity of 0.90, the specificity was 0.902. In each participating center, AUROC ranged from 0.913 to 0.970 (Supplementary Figure 3). When restricted the analysis in either patients without LVO or isolated MCA-M2 occlusion, AUROC was 0.880 (95% CI, 0.824 – 0.921, Figure 1B). At the highest Youden index (0.657; 95% CI, 0.514 – 0.764), the optimal criterion, sensitivity, and specificity were 0.405, 0.763, and 0.894, respectively.

**Figure 1.**
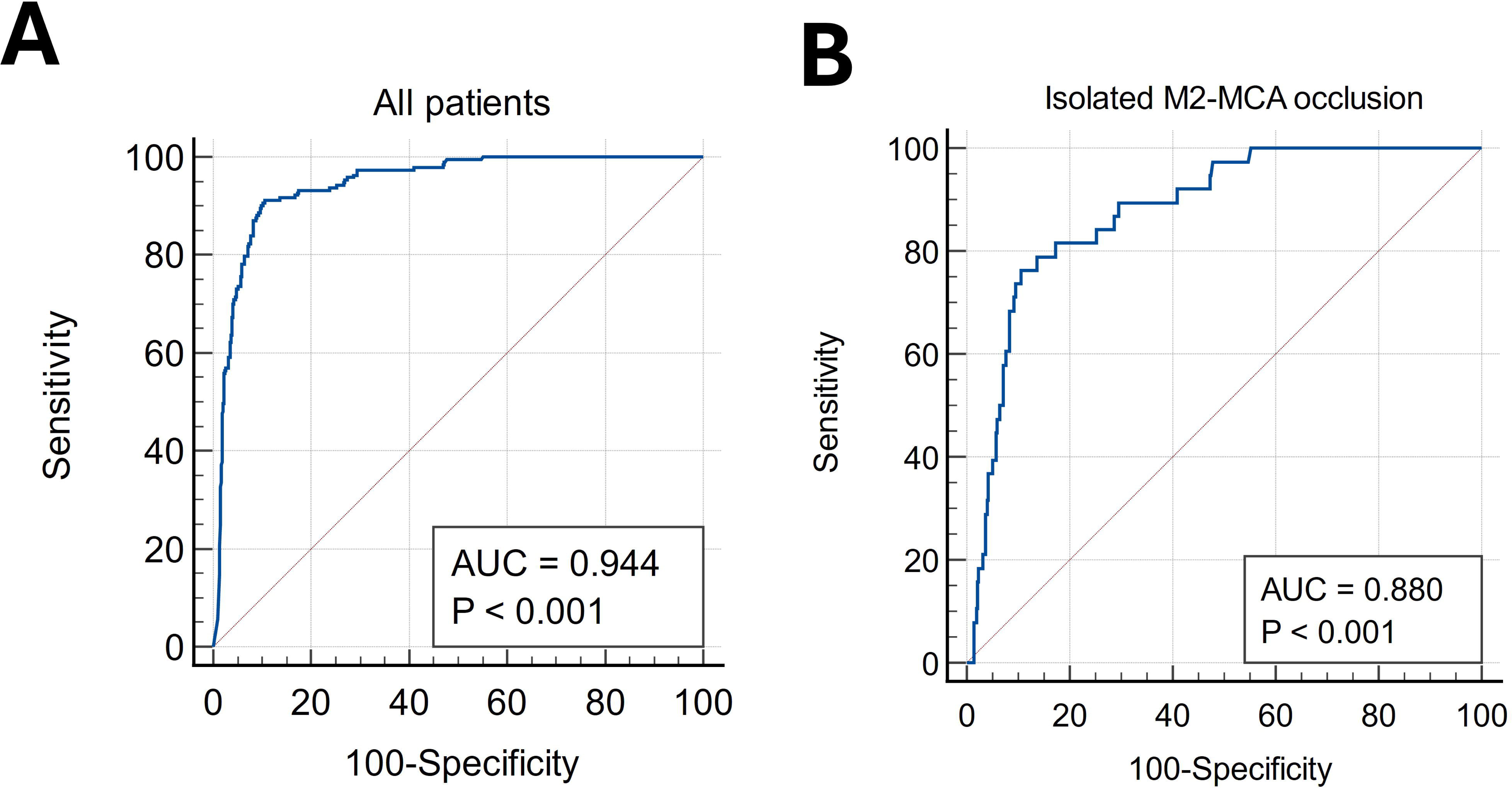
Diagnostic performance of JLK-LVO in the entire population. (A) Overall diagnostic performance, (B) diagnostic performance in patients with isolated MCA-M2 occlusion

**Table 2.**
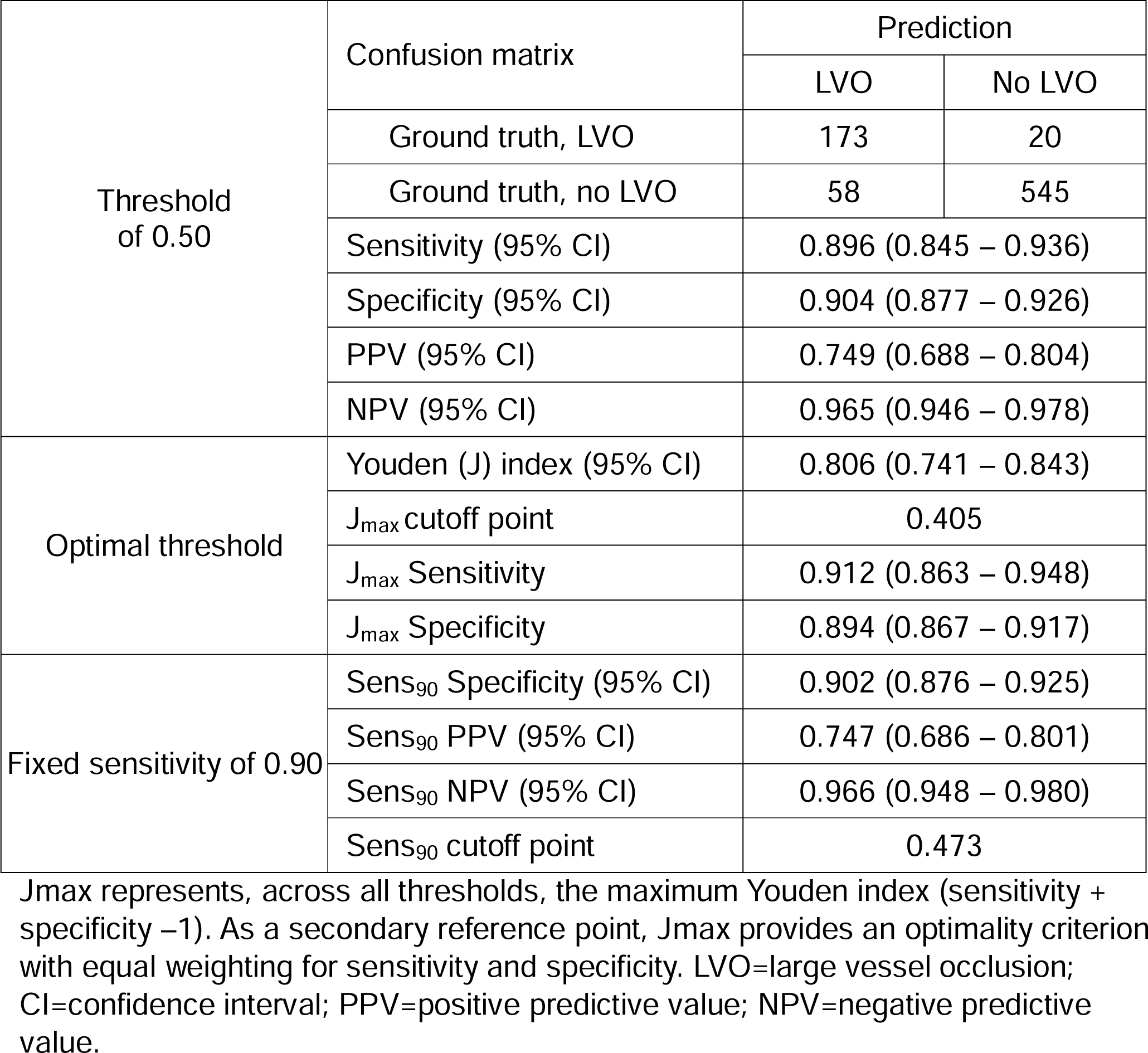
Diagnostic performance of software detecting large vessel occlusion.

### LVO scores according to vessel status

When stratified patients into five distinct groups (acute LVO, chronic LVO, isolated MCA-M2 occlusion, relevant MCA stenosis, and neither stenosis nor occlusion of MCA), the medians (IOR) of LVO scores were 99.8 (97.2 – 99.97), 99.1 (97.3 – 99.99), 82.1 (40.9 – 98.2), 15.3 (2.4 – 77.4), and 0.5 (0.1 – 6.5), respectively (Figure 2). Compared with the neither stenosis nor occlusion of MCA groups, the median LVO scores of relevant MCA stenosis group was significantly higher (p < 0.001).

**Figure 2.**
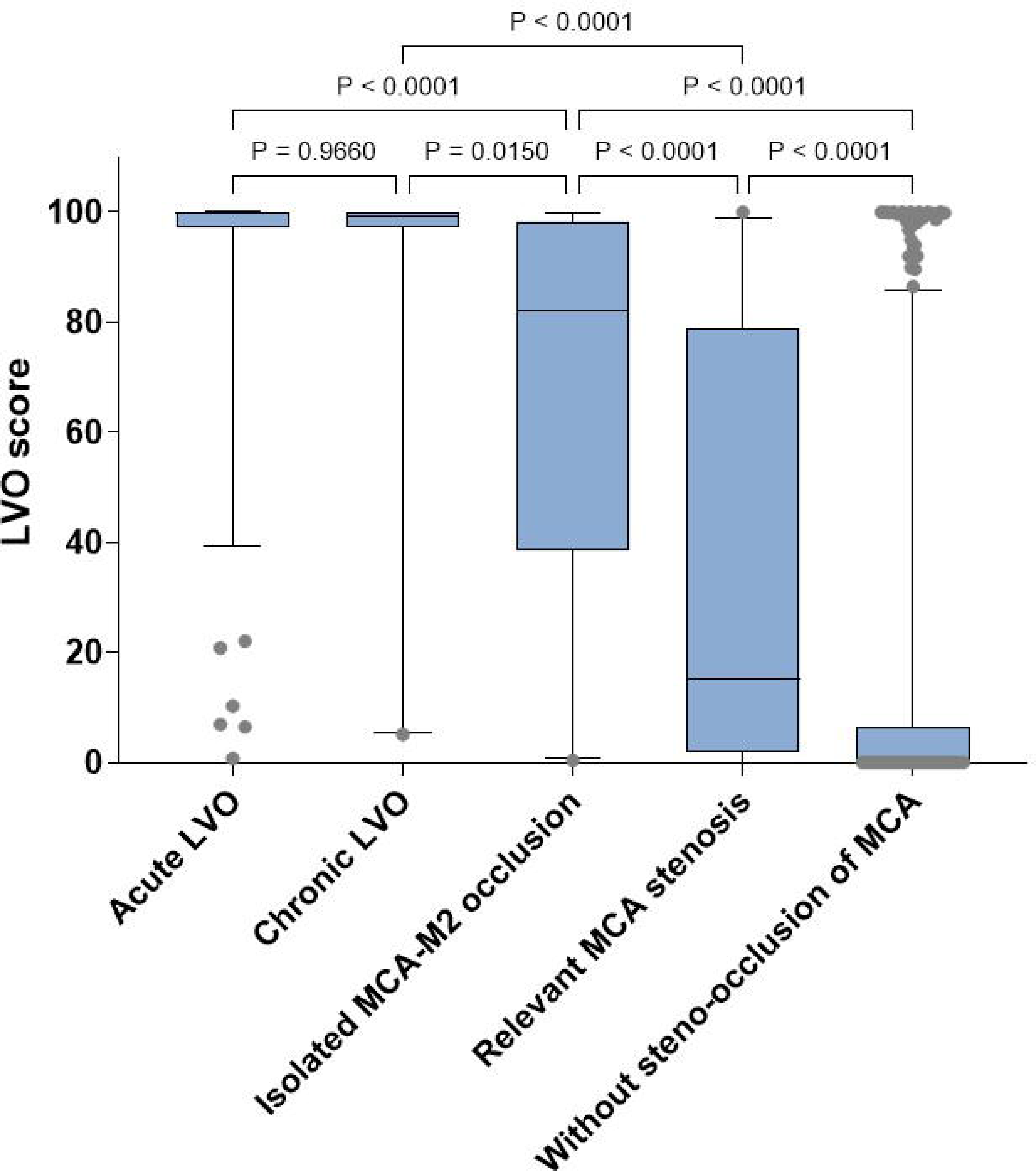
Box plots for LVO score s after stratifying patients according to vessel status. LVO = large vessel occlusion; MCA=middle cerebral artery. Boxes and midline indicate interquartile ranges and the median of LVO scores. Whiskers indicate 5∼95 percentile of data.

### Calibration of LVO score

Before calibration, unadjusted LVO probabilities significantly overestimated LVO, as the observed/expected LVO ratio was 0.792 (Figure 3A). After calibration, the adjusted LVO probabilities achieved an observed/expected LVO ratio of 1.00 (Figure 3B). However, due to sparse data between LVO probabilities of 0.2 to 0.6, the point estimations at adjusted probabilities of 0.4, 0.6, and 0.8 exhibited discrepancies between the expected and observed frequencies of LVO.

**Figure 3.**
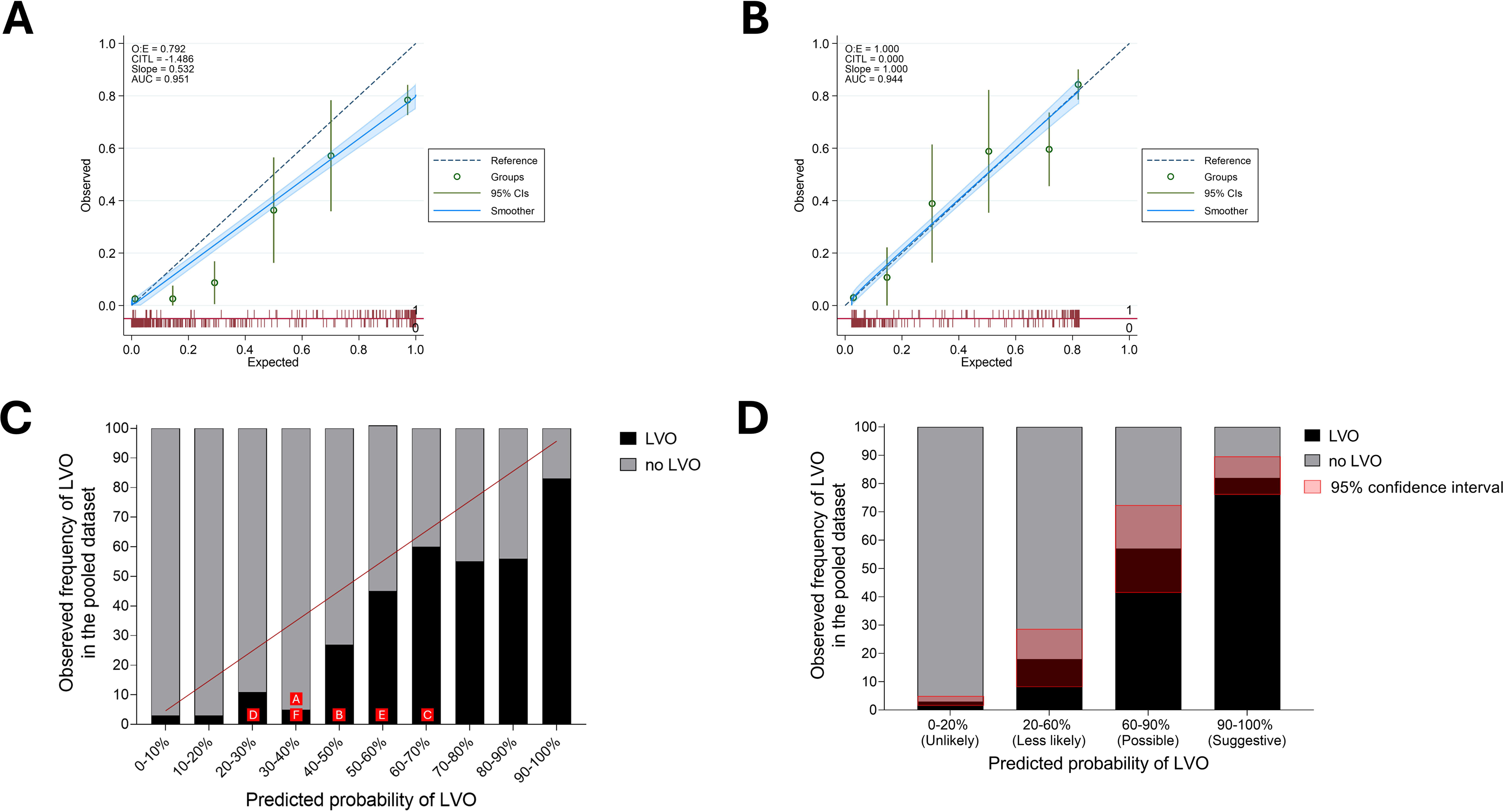
Predicted frequency and observed frequency of LVO before and after calibration of LVO score. (A, B) A calibration plot showing observed probability against expected probability using either unadjusted LVO probability (A) and adjusted LVO probability (B). The green dotted line indicates the reference line of perfect agreement. Red spikes indicate each case with LVO (up spike) without LVO (down spike) at each LVO probability. O:E=ratio of observed and expected LVO frequency; CITL= Calibration-in-the-large, also known as mean calibration. (A) (C) The red line indicates perfect calibration. The red boxes indicate the criterion with the highest Youden index in each hospital. (D) Observed frequencies and their 95% confidence intervals (red shaded areas) after recategorizing of patients. LVO=large vessel occlusion

As the LVO score percentile increased, the observed frequency of LVO increased in a stepwise manner (p for trend < 0.001; Figure 3C). However, due to the bimodal distribution of LVO scores (Supplementary Figure 2), the observed frequencies of LVO between 20% and 90% showed low concordance compared to the predicted probability. Based on these results, we recategorized subjects into four groups: unlikely (0-20% LVO scores), less likely (20-60%), possible (60-90%), and suggestive (90-100%). After recategorization, each group represented observed frequencies well without overlapping confidence intervals (Figure 3D). In addition, observed frequencies of EVT were 5, 10, 24, and 49% in each group, respectively (Supplementary Figure 4).

### Associations of LVO scores with infarct volumes and functional outcome

Follow-up DWIs within 7 days of the last known well were available for 711 (89.3%) patients. The median (IQR) interval between CTA and DWI was 1.5 (0.8 – 2.8) hours. The median (IQR) infarct volumes of the unlikely, less likely, possible, and suggestive groups were 1.1 mL (0.2 – 6.4 mL), 4.4 mL (0.5 – 24.4 mL), 6.2 mL (0.6 – 38.6 mL), and 20.6 mL (2.5 – 104.5 mL), respectively (p for difference < 0.001; Figure 4A). Additionally, we observed a significant trend of shifting 3-month modified Rankin Scale scores to higher scores as the recategorized LVO score groups increased (p < 0.001; see Figure 4B).

**Figure 4.**
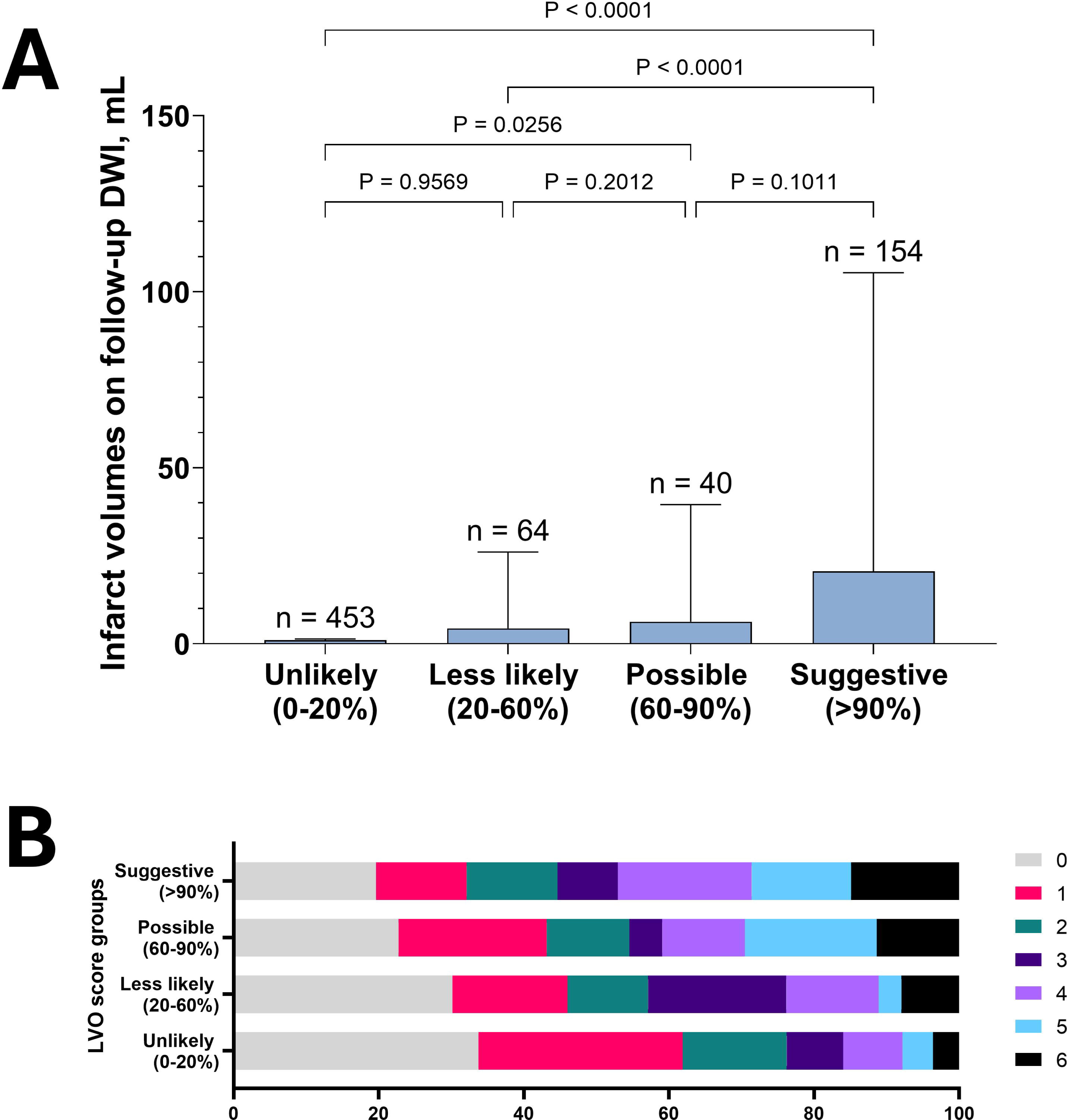
Relation of LVO scores with follow-up infarct volume and functional outcome. (A) Bars and error bars represent the median and its 95% confidence interval. For post-hoc comparison, the Tukey method was used. (B) Distributions of 3-month modified Rankin Scale scores according to LVO score gorups.

## Discussion

In this multicenter study comprising 796 consecutive series of patients with ischemic stroke or transient ischemic attack from 6 university hospitals, we observed the robust clinical efficacy of JLK-LVO, an automated software detecting LVO on CTA utilizing a deep learning algorithm. Using a real-world clinical dataset, we calibrated the LVO score derived from deep learning and suggested a new category for better understanding of probability for clinicians. Additionally, we found associations of the new category of LVO score with infarct volumes on follow-up DWI and 3-month modified Rankin Scale scores.

Using a multicenter dataset with various CT vendors and imaging parameters, JLK-LVO exhibited robust and consistent AUROC values ranging from 0.918 to 0.970. The deep learning algorithm in this study was trained on a large dataset of over 2,700 CTA scans from five hospitals,^13^ enabling it to maintain its performance across different datasets and potentially enhancing its generalizability without further training. Additionally, JLK-LVO achieved a sensitivity of 76% in detecting isolated MCA-M2 occlusion, which is even higher than that reported by neuroradiologists in a study involving 520 patients with ischemic stroke; experienced neuroradiologists missed 35% of MCA-M2 occlusions during initial CTA evaluation.^20^ Given the recent efforts to expand EVT candidacy to MCA-M2 segment occlusions,^21^ the ability to detect MCA-M2 occlusions with high accuracy may facilitate the treatment and benefit of more patients undergoing EVT.

In medical contexts, there is often an imbalance between normal and abnormal data, which hampers deep learning model training.^22^ Data augmentation^23^ and random under-sampling^24^ are common techniques for addressing class imbalance, often improving model performance. However, in cases where augmentation may distort data, model calibration may be considered to compensate for the imbalance. Ensuring effective confidence calibration for deep learning models enhances the reliability of their predictions, which is crucial for their practical deployment in safety-critical tasks like medical diagnosis.^25^ In the present study, we observed a notable bimodal distribution of LVO probabilities score, which, in turn, renders calibration challenging in the range with scarce data. Additionally, different optimal criteria across participating centers suggest that a model-based calibration, commonly used in deep learning algorithms,^26^ is less practical and prone to miscalibration due to the highly variable disease prevalence and imaging parameters in clinical practice. Hence, we collapsed multiple categories with a similar observed frequency of LVO into one and generated four groups that distinctly represent the observed frequency of LVO. We believe that this calibrated interpretation, along with uncertainty (the range of observed frequency), provides more reliable results for clinicians.

Of note, LVO scores in patients with relevant MCA stenosis were significantly higher (median 15.3 vs. 0.5) compared to those without MCA stenosis or occlusion. This result indicates that the deep learning algorithm utilizes the symmetry of vascular density between hemispheres as an important feature to detect LVO. Consistent with this finding, we observed an association of LVO score groups with infarct volume on follow-up DWI and 3-month modified Rankin Scale score. If confirmed in future studies, the LVO score derived from deep learning may be utilized as a novel biomarker to select patients who may potentially benefit from EVT.

The large size of a consecutive series of CTA data from various vendors and imaging parameters is a strength of our study. Nevertheless, several limitations should be acknowledged. We collected data taken in the emergency room from university hospitals. Hence, further study is required to extrapolate our results to outpatient settings or community hospitals. Additionally, the different head sizes and nature of LVO across ethnicities limited the generalizability of our results. Ongoing trials involving other ethnic groups may provide insights into this issue.

In conclusion, this multicenter study confirmed the clinical efficacy of deep learning algorithm (JLK-LVO, JLK Inc., Seoul, Korea) in various CT vendors and imaging parameters. Robust performance of the algorithm along with high accuracy in MCA-M2 occlusion detection may facilitate stroke workflow particularly in the community with limited resources.

## Data Availability

All data produced in the present study are available upon reasonable request to the authors.

