## Supplementary Material for "Automated detection of large vessel occlusion: a multicenter study validating efficacy and proving clinical implications"

**Supplementary Figure 1. Study flow chart**

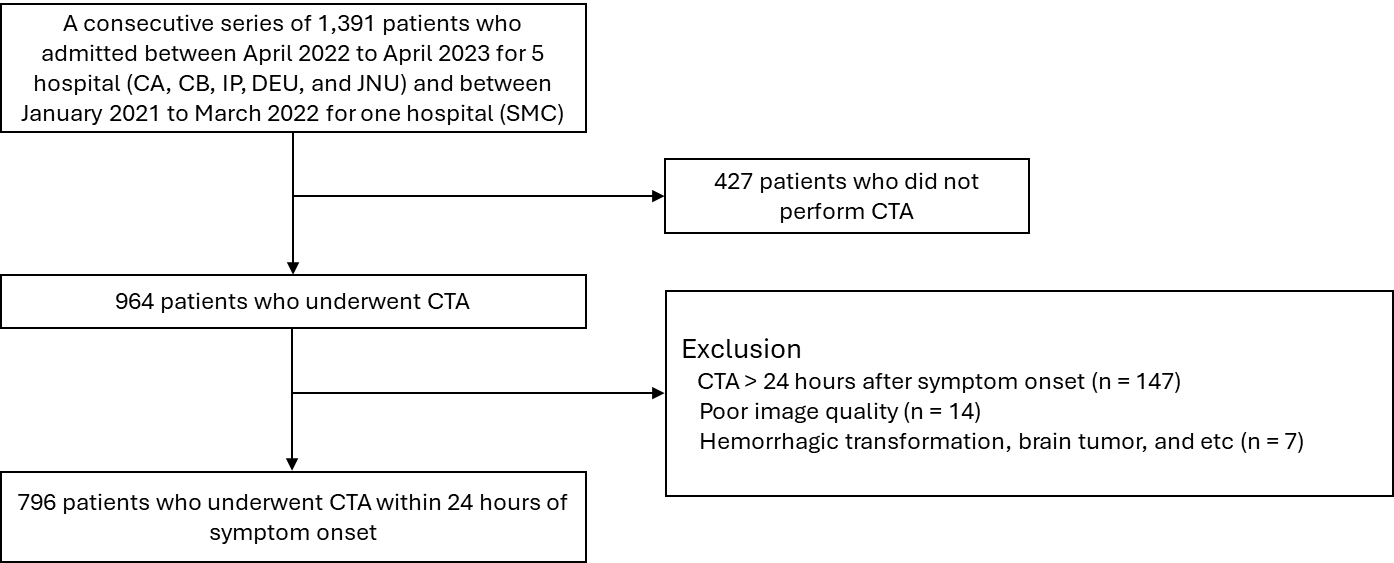

**Supplementary Figure 2. Histogram of LVO score according to the presence of LVO**

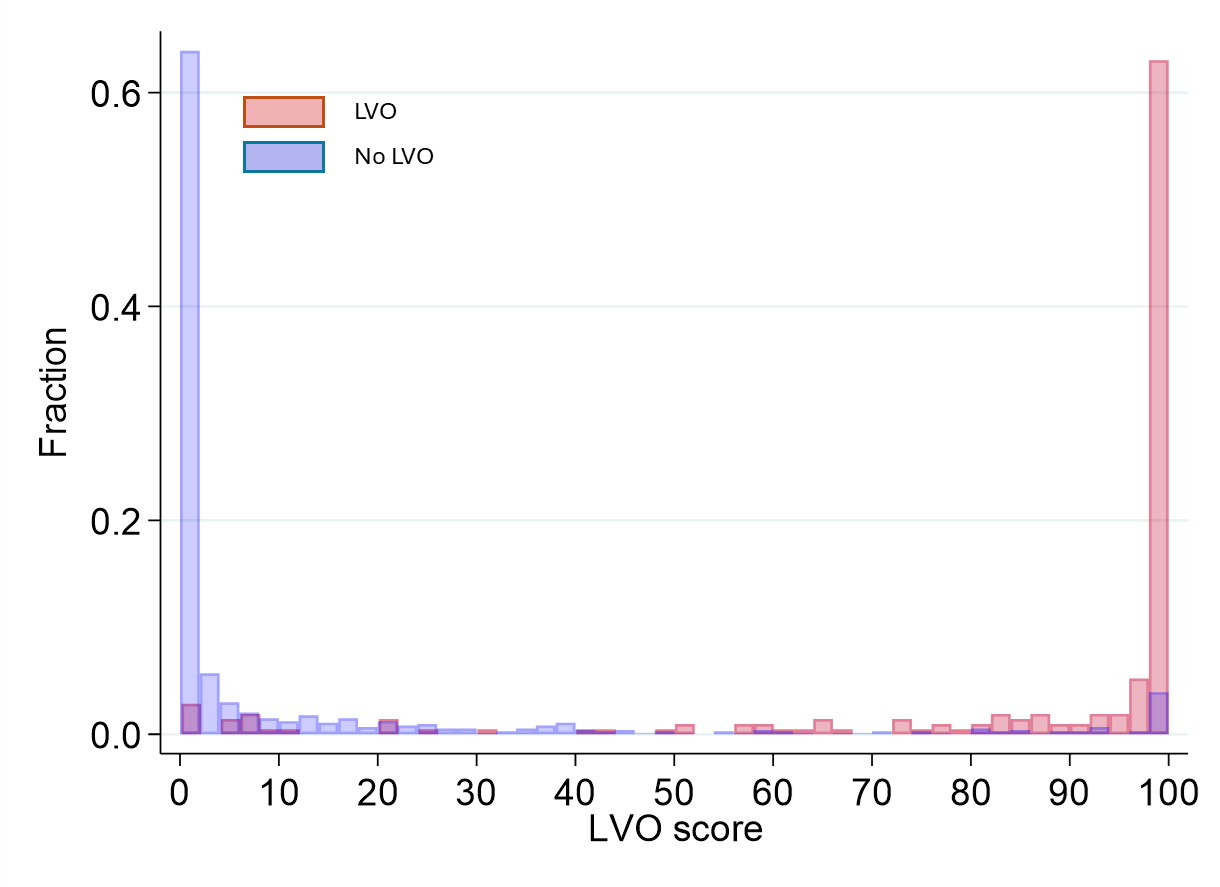

**Supplementary Figure 3. Area under the receiver operating characteristics curve in each participating center**

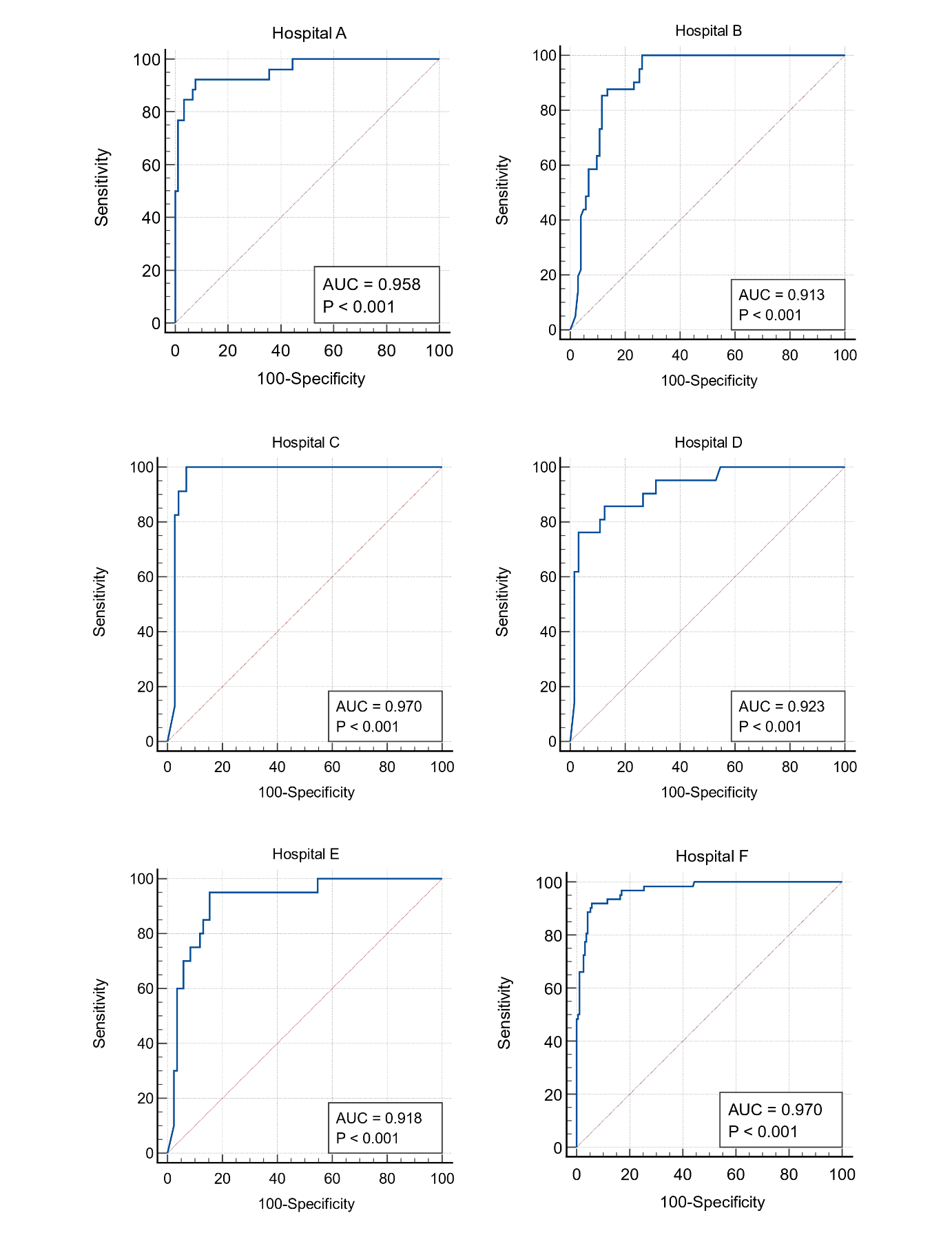

**Supplementary Figure 4. Observed frequency of endovascular treatment across LVO score groups**

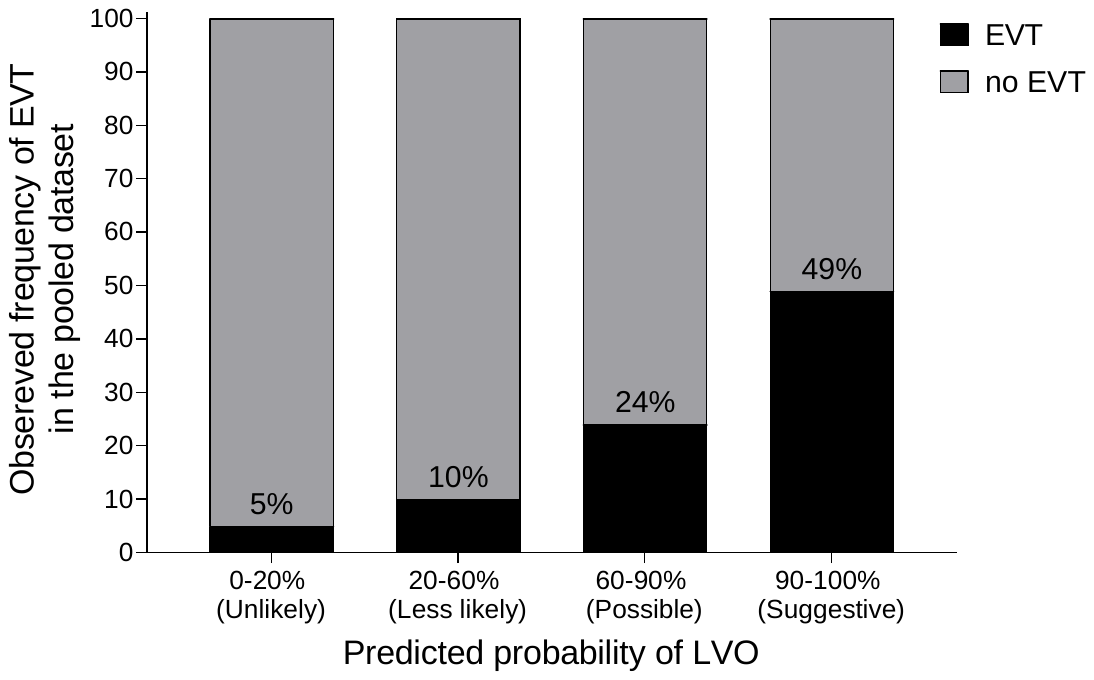

EVT=endovascular treatment; LVO=large vessel occlusion**.**

**Supplementary table 1. CT angiography parameters in participating centers**

|  | Hospital A  (n = 116) | Hospital B  (n = 144) | Hospital C  (n = 96) | Hospital D  (n = 85) | Hospital E  (n = 104) | Hospital F  (n = 251) |
| --- | --- | --- | --- | --- | --- | --- |
| Slice thickness, mm | 1.5 | 0.625 | 0.75 | 1.0 | 3 | 1.25 |
| CT manufacture | Phillips | GE medical systems | SIEMENS | SIEMENS | TOSHIBA | GE medical systems |
| CT model name | iCT 256 | Revolution CT | SOMATOM Definition AS+, SOMATOM Definition Flash | SOMATOM Definition Edge, SOMATOM Force | Aquilion PRIME | Discovery CT750 HD |
| kVp | 120 | 120 | 120 | 90 (n = 28)  100 (n = 3)  120 (n = 54) | 120 | 120 |
| Rotation time,^a^ s | 0.33 | 0.5 | NA | NA | 0.5 | 0.6 |
| Total Collimation Width, mm | 40 | 40 | 38.4 | 38.4 | 40 | 40 |
| Spiral Pitch Factor | 0.515 (n = 62)  0.601 (n = 54) | 0.984375 | 1.2 | 0.45 (n = 26)  0.7 (n = 28)  1.0 (n = 27) | 0.813 | 0.984375 |
| mAs | 140 (n = 54)  200 (n = 62) | 249.5 ~ 251.5 | 327 ~ 338.5 | 61.75 ~ 330 | 100 (n = 2)  125 (n = 102) | 60 ~ 376.2 |

^a^Rotation time was not available in SIEMENTS CT scanners.
